# Comparative effectiveness of different primary vaccination courses on mRNA based booster vaccines against SARs-COV-2 infections: A time-varying cohort analysis using trial emulation in the Virus Watch community cohort

**DOI:** 10.1101/2022.02.04.22270479

**Authors:** Vincent Grigori Nguyen, Alexei Yavlinsky, Sarah Beale, Susan Hoskins, Vasileios Lampos, Isobel Braithwaite, Thomas E Byrne, Wing Lam Erica Fong, Ellen Fragaszy, Cyril Geismar, Jana Kovar, Annalan M D Navaratnam, Parth Patel, Madhumita Shrotri, Sophie Weber, Andrew C Hayward, Robert W Aldridge, the Virus Watch Collaborative

## Abstract

**Importance:** The Omicron (B.1.1.529) variant has increased SARs-CoV-2 infections in double vaccinated individuals globally, particularly in ChAdOx1 recipients. To tackle rising infections, the UK accelerated booster vaccination programmes used mRNA vaccines irrespective of an individual’s primary course vaccine type with booster doses rolled out according to clinical priority. There is limited understanding of the effectiveness of different primary vaccination courses on mRNA based booster vaccines against SARs-COV-2 infections and how time-varying confounders can impact the evaluations comparing different vaccines as primary courses for mRNA boosters.

**Objective:** To evaluate the comparative effectiveness of ChAdOx1 versus BNT162b2 as primary doses against SARs-CoV-2 in booster vaccine recipients whilst accounting for time-varying confounders.

**Design:** Trial emulation was used to reduce time-varying confounding-by-indication driven by prioritising booster vaccines based upon age, vulnerability and exposure status e.g. healthcare worker. Trial emulation was conducted by meta-analysing eight cohort results whose booster vaccinations were staggered between 16/09/2021 to 05/01/2022 and followed until 23/01/2022. Time from booster vaccination until SARS-CoV-2 infection, loss of follow-up or end-of-study was modelled using Cox proportional hazards models for each cohort and adjusted for age, sex, minority ethnic status, clinically vulnerability, and deprivation.

**Setting:** Prospective observational study using the Virus Watch community cohort in England and Wales.

**Participants:** People over the age of 18 years who had their booster vaccination between 16/09/2021 to 05/01/2022 without prior natural immunity.

**Exposures:** ChAdOx1 versus BNT162b2 as a primary dose, and an mRNA booster vaccine.

**Results:** Across eight cohorts, 19,692 mRNA vaccine boosted participants were analysed with 12,036 ChAdOx1 and 7,656 BNT162b2 primary courses with a median follow-up time of 73 days (IQR:54-90). Median age, clinical vulnerability status and infection rates fluctuate through time. 7.2% (n=864) of boosted adults with ChAdOx1 primary course experienced a SARS-CoV-2 infection compared to 7.6% (n=582) of those with BNT162b2 primary course during follow-up. The pooled adjusted hazard ratio was 0.99 [95%CI:0.88-1.11], demonstrating no difference between the incidence of SARs-CoV-2 infections based upon the primary vaccine course.

**Conclusion and Relevance:** In mRNA boosted individuals, we found no difference in protection comparing those with a primary course of BNT162b2 to those with aChAdOx1 primary course. This contrasts with pre-booster findings where previous research shows greater effectiveness of BNT162b2 than ChAdOx1 in preventing infection.

## Introduction

England and Wales has recently experienced an increase in SARs-CoV-2 infections in individuals that received two vaccines. This increase in infection rates is partially attributable to waning vaccine protection and the emergence of the variant of concern, Omicron (B.1.1.529) which has mutations leading to partial immune escape from prior infection or vaccination. To tackle the growth in infections, the UK accelerated booster vaccines to those who received two doses of certified COVID-19 vaccinations with a gap of 3-months between the second and a third booster dose.

Our previous analysis found a difference in SARs-CoV-2 infection rates between the two dominant vaccines (ChAdOx1 or BNT162b2) in the United Kingdom with those receiving ChAdOx1 as their primary course having a 35% increased risk of having a SARs-CoV-2 infection 1.35 [HR: 1.35, 95%CI: 1.15 - 1.58] at up to 315 days post first vaccination ^1^. Our findings are consistent with previous work which demonstrated the difference in peak Spike-antibody levels (the primary antibody stimulated by vaccination-related inoculation) based upon vaccine type; where BNT162b2 produced Spike-antibody levels an order of magnitude higher than ChAdOx1 after two doses ^2(p1)^. Due to the difference in vaccine effectiveness in preventing SARs-CoV-2 infections and enhancing antibody levels, and data from recently conducted randomised controlled trials examining safety and immunogenicity of seven COVID-19 vaccines as a booster dose^3^, messenger ribonucleic acid (mRNA) based vaccines (BNT162b2 or mRNA-1273) were chosen as viable options for the booster dose in the UK to tackle further waves of infection^3^.

Following the use of BNT162b2 or mRNA-1273 as booster doses in the UK, research from the UK Health Security Agency (UKHSA) has demonstrated similar effectiveness between primary vaccine courses and mRNA boosters using test-negative study designs ^4^. Test negative designs are well suited to reducing biases related to test seeking behaviour ^5^ but are subject to temporal confounding where timing of vaccination is influenced by risk factors for infection ^6^ To protect the most vulnerable and exposed to SARs-CoV-2, the UK’s strategy prioritised booster vaccination roll out based upon age, clinical vulnerability and exposure to the virus (for example, frontline healthcare workers). In addition to variation in timing of booster vaccinations according to risk factors, there are substantial variations in levels of infection and intensity of control measures over time which complicate the task of controlling for time varying confounding.

In this study, we aim to apply trial emulation techniques developed by Hernan and Robins ^7^ to tackle time-varying confounding by indication. Following Hernan et al’s recommendations to overcome time varying confounding we use an eligibility criteria that removes those who are likely to have protection from SARs-COV-2 (e.g. through prior natural infection) and stagger our cohort based upon vaccination date. Staggering a single cohort into multiple cohorts aims to produce cohorts that are homogenous in terms of the eligibility criteria that had allowed them to be vaccinated at that point in time. Using staggered cohorts also allows similar individuals to have similar followup periods and more importantly, experience the same COVID-19 public health policies and SARs-CoV-2 reproduction rates at the time of vaccination and throughout their followup period. This approach aims to control for the UK’s booster prioritization list but could also mitigate the effects of unmeasured time varying confounders at the community level, for example, the introduction of new SARS-CoV-2 variants. Therefore, this approach appropriately accounts for “time zero” (start of followup) as it avoids comparisons between individuals who experienced different public health policies and SARs-CoV-2 reproduction rates through time.

In this study, our objective is to use a trial emulation approach to appropriately estimate the comparative effectiveness of receiving different primary vaccine courses (ChAdOx1 or BNT162b2) in addition to an mRNA booster vaccine against SARs-CoV-2 infections in a general population community cohort.

## Method

### Study Design and Setting

The study design used prospective observational data from the Virus Watch Cohort and applied a target trial emulation study design - a detailed description of the target trial emulation can be found in Table 1. The Virus Watch cohort has been described previously ^8^. Briefly, households were recruited starting in mid-June 2020 via several methods aimed at creating a representative cohort of England and Wales, including postcards sent to the home address, social media, and SMS. As of January 2022, 58,634 individuals in 28,525 households had registered to take part. Participants completed weekly online surveys reporting symptoms, SARS-CoV-2 swab test results and vaccinations. From Autumn 2020, Virus Watch also included a programme of nasopharyngeal swab sample collection and blood collection via venipuncture or finger prick sampling in a subset of 10,000 participants in research clinics. From March 2021, blood samples were self-collected by participants using an at-home capillary blood sample collection kit, manufactured by the company Thriva [https://thriva.co/]. Completed kits were returned by participants using pre-paid envelopes and priority postage boxes to UKAS-accredited laboratories for serological testing using Roche’s Elecsys Anti-SARS-CoV-2 assays targeting total immunoglobulin (Ig) to the Nucleocapsid (N) protein or to the receptor-binding domain in the S1 subunit of the Spike protein (S) (Roche Diagnostics, Basel, Switzerland).

**Table 1:** Details of the trial emulation framework used to conceptualise the observational study as a controlled trial

|  | **Ideal Randomised Controlled Trial** | **Trial emulation** |
| --- | --- | --- |
| **Eligibility Criteria** | -At least 18 years old<br>-No prior SARS-CoV-2 infection<br>-Two doses of the SARs-CoV-2 vaccine | -At least 18 years old when vaccinated<br>-No recorded SARS-CoV-2 infection prior to booster vaccination date, defined using: PCR, LFT, nucleocapsid antibodies and spike protein before 2021<br>-Two doses of the SARs-CoV-2 vaccine |
| **Recruitment period** | September 16th 2021 to January 05 2022 | September 16th 2021 to January 05 2022 split by 14-day intervals:<br>-Cohort 1: 2021-09-16 to 2021-09-29<br>-Cohort 2: 2021-09-30 to 2021-10-13<br>-Cohort 3: 2021-10-14 to 2021-10-27<br>-Cohort 4: 2021-10-28 to 2021-11-10<br>-Cohort 5: 2021-11-11 to 2021-11-24<br>-Cohort 6: 2021-11-25 to 2021-12-08<br>-Cohort 7: 2021-12-09 to 2021-12-22<br>-Cohort 8: 2021-12-23 to 2022-01-05 |
| **Follow-up duration** | From 2021-09-16 to 2022-01-23 | From recorded booster vaccination date until 2022-01-23 |
| **Outcome** | 1) Positive PCR test for SARS-CoV-2<br>2) Positive LFT for SARS-CoV-2 | 1) Positive PCR test for SARS-CoV-2 (self-reported or linked data )<br>2) Positive LFT for SARS-CoV-2 (self-reported or linked data) |
| **Treatments to be compared** | -Booster Dose with primary course of ChAdOx1<br>-Booster Dose with a primary course of BNT162b2 | -Booster Dose with primary course of ChAdOx1<br>-Booster Dose with a primary course of BNT162b2 |
| **Estimand** | Intention to treat based upon primary course | Intention to treat based upon primary course |
| **Analysis plan** | Survival analysis (Kaplan Meier estimator) | Survival analysis (pooled multivariable Cox Proportional hazard models) |

### Participants

Participants from the Virus Watch cohort were eligible for the current analysis if they received a third (booster) COVID-19 vaccination recorded between September 16^th^ 2021 and January 5^th^ 2022. Participants must have had a primary COVID-19 vaccination dose recorded of either ChAdOx1 or BNT162b2. As the UK vaccination programme only included children under 18 years of age in the second half of 2021, participants under 18 years old were excluded from these analyses due to their low numbers. Participants who had evidence of SARS-CoV-2 infection prior to their booster vaccination were excluded to examine vaccine and not natural infection-related immunity. Previous SARS-CoV-2 infection was defined using the following: 1) A positive self-reported PCR or LFT test, 2) a positive PCR or LFT test from data linkage 3) presence of IgN through venous sampling or 4) the presence of the Spike antibody prior to December 2020 as these were likely due to natural infection or participation in a vaccination trial.

### Exposure Variables

The exposure variable was the vaccination type (ChAdOx1 or BNT162b2) of the primary vaccine course. Vaccination data in Virus Watch was combined from self-reported and linked data from the National Immunisation Management Service (*NIMS*) dataset.

In the January 11^th^, 2021 and January 18th, 2021 Virus Watch questionnaires, participants were asked about their vaccination status retrospectively. From 25 January 2021 onwards, participants were asked weekly for their vaccination status. Recorded vaccinations from NIMS covered the period October 9th, 2020, until December 23^rd^ 2021.

### Outcome Variables

The primary outcome was SARS-CoV-2 infection using: 1) a positive self-reported PCR or LFT test or 2) a positive PCR or LFT test from the linked Second-Generation Surveillance System (SGSS) data. As we did not link our data on points 1 or 2 to symptom data, and our outcome may therefore include asymptomatic cases, we refer to our primary outcome as SARS-CoV-2 infection rather than COVID-19 disease for the purposes of this analysis, although as most testing is undertaken in response to symptoms the cases will largely represent symptomatic rather than asymptomatic infection.

### Covariates

Self-reported demographic data included age, sex, and ethnicity. We included clinically vulnerable status which was derived from self-reported data on immunosuppressive therapy, cancer diagnoses, and chronic disease status. Index of multiple deprivation quintiles was derived based upon Lower Layer Super Output Areas postcodes submitted during registration. Due to small sample sizes (particularly by staggering cohorts), we could not evaluate geographical region or ethnicity in detail; therefore, we classified ethnicity as “White British” or “Ethnic Minority”.

### Data Sources and Linkage

For SARs-CoV-2 infections, the primary source of data was the Virus Watch dataset linked to the Second-Generation Surveillance System (SGSS), which contains SARS-CoV-2 test results using data from hospitalisations (Pillar 1) and community testing (Pillar 2). Linkage was conducted by NHS Digital with the linkage variables being sent in March 2021. The linkage period for SGSS Pillar 1 encompassed data from March 2020 until August 2021 and from June 2020 until November 2021 for Pillar 2.

For vaccination data, the primary source of data was the Virus Watch dataset linked to the National Immunisation Management Service (*NIMS*) and encompasses vaccinations between October 9^th^ 2020 and December 23^rd^ 2021.

### Bias

To estimate the risk of SARs-CoV-2 infection after receiving the booster COVID-19 vaccine, time-to-event analyses could be conducted. However, evaluating time to SARs-CoV-2 infection may be confounded by the United Kingdom’s strategy to prioritise booster vaccinations based upon age, clinical vulnerability, and exposure to the virus (for example, frontline healthcare workers). To tackle such biases, we used methods developed by Hernan and Robins that aim to tackle confounding by indication to appropriately estimate the average treatment effect ^7,9^. This approach includes three primary components 1) excluding prevalent users of an intervention to estimate the impact of treatment initiation without the lingering effect of previous confounding treatment, 2) use of an intention to treat analysis as this is the common estimand in randomised controlled trials, and 3) the use of multiple staggered cohorts to appropriately account for “time zero” (or the start of follow-up).

To apply Hernan et al.’s recommendations, this study 1) use eligibility criteria that exclude those who are likely to have alternative protection from SARs-COV-2 (e.g., through prior natural infection), 2) assign individuals from the first vaccination dose and disregard changes in course and 3) stagger a single cohort based upon vaccination date; see the introduction for rationale of cohort staggering.

### Statistical Analysis

Pooled Cox proportional hazard models were used to estimate the time from vaccination until the primary outcome of SARS-CoV-2 infection, lost to follow-up (latest week of reporting to Virus Watch), or end of study (January 23^rd^ 2022), whichever was earliest. Cohorts were split based upon the date of their booster vaccination with the cohort dates defined in table 1.

Multivariable adjustment was conducted using the following variables: age, sex, ethnic minority status, index of multiple deprivation quintiles and clinical vulnerable status (clinically vulnerable, clinically extremely vulnerable, or none identified). The models from the eight cohorts were pooled using a random-effects meta-analysis. Full case analysis was conducted for all analyses. Statistical analysis was conducted using R version 4.0.3,

### Ethical approval

This study has been approved by the Hampstead NHS Health Research Authority Ethics Committee. Ethics approval number - 20/HRA/2320.

## Results

Across the eight cohorts, among those who met the eligibility criteria (adults, with a recording of ChAdOx1 or BNT162b2 as a primary course without prior infection or missing data), a total of 19,692 participants received their booster vaccination between September 16th 2021 to January 5th 2022. The largest recruitment period was between October 28th 2021 to November 10th 2021, with 4,558 participants. The smallest recruitment period was between December 23rd 2021 to January 05 2022, with 350 participants. See Supplementary Figure 1 for the recruitment timeline of the emulated trials.Across all eight cohorts, 12,036 individuals had received the ChAdOx1 vaccine, whilst 7,656 received the BNT162b2 vaccine. Demographic characteristics were broadly similar between ChAdOx1 and BNT162b2, except for clinical vulnerability status and age, where BNT162b2 had slightly more clinically vulnerable patients with an older age group (See Table 2). Due to the staggered cohort design, it is more appropriate to compare individuals who were vaccinated in the same period. In brief, as time advanced, recipients of the booster doses were getting younger; prior to mid-December 2021, the age distributions for both vaccines were similar, however, after this period, those whose primary course was BNT162b2 were younger than their ChAdOx1 counterparts for any given day (Figure 1a). In terms of clinically vulnerability, both ChAdOx1 and BNT162b2 saw a decrease in the daily proportion of those identified as “clinically extremely vulnerable”(Figure 1b)

**Figure 1a (top).**
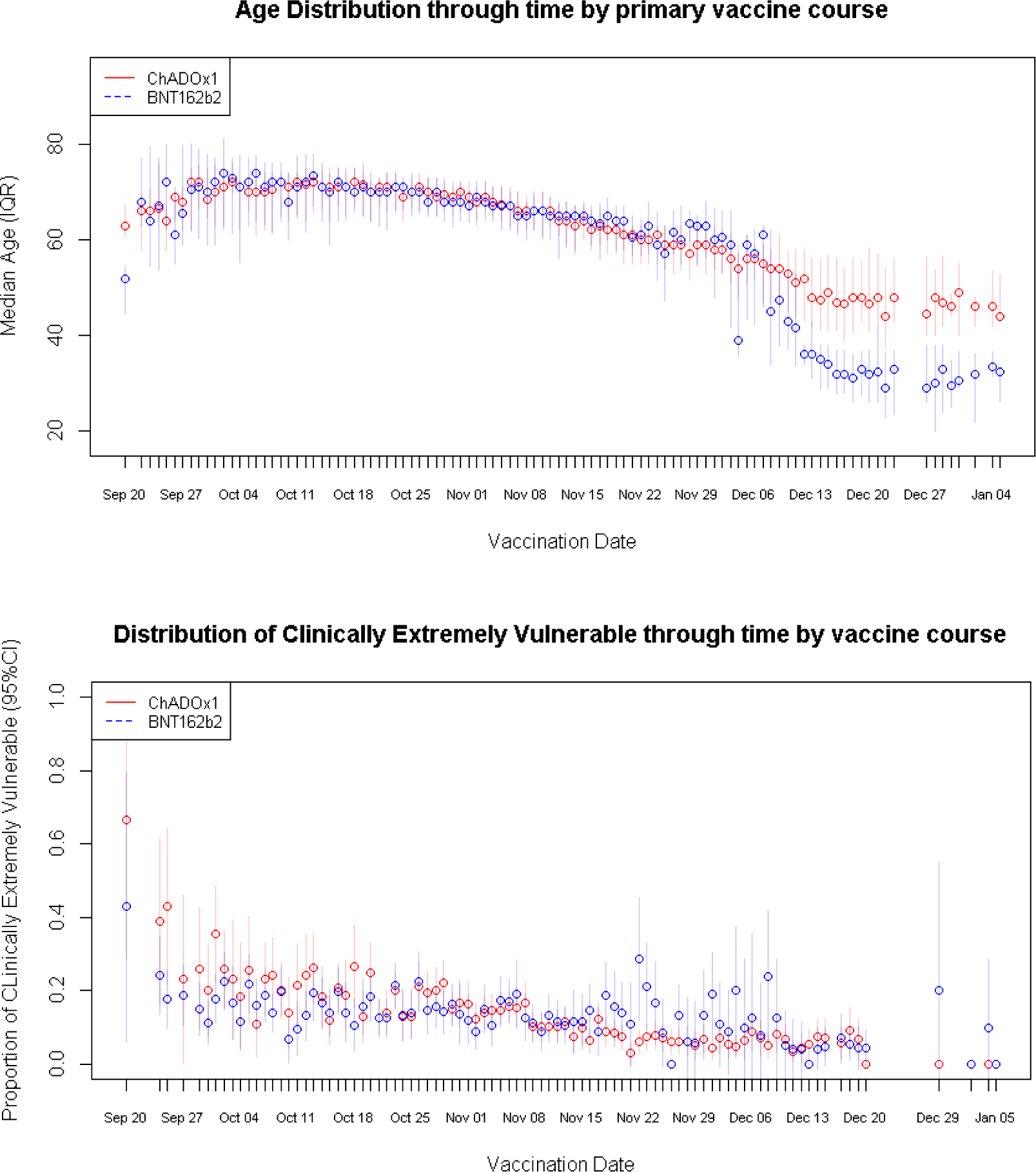
Distribution of age through time by primary vaccination course. The gradient for ChAdOx1 was decrease of 0.27 years per day [95%CI: −0.29 to −0.24] whilst the gradient for BNT162b2 was also a decrease 0.41 years per day [95%CI: −0.46 to −0.35]. Figure: 1b (bottom): Distribution of extremely clinical vulnerable status through time by primary vaccination course. The gradient for ChAdOx1 was a daily decrease of 0.003 proportions of Clinically Extremely Vulnerable [95%CI: −0.003 to −0.002] whilst the gradient for BNT162b2 was a daily decrease proportion of Clinically Extremely Vulnerable 0.001[95%CI: −0.001 to −0.001].

**Table 2:** Demographics breakdown of the cohort.

| Characteristic | N | BNT162b2 ,<br>N = 7,656<sup>1</sup> | ChAdOx1 ,<br>N = 12,036<sup>1</sup> | p-value<sup>2</sup> |
| --- | --- | --- | --- | --- |
| **Age** | 19,692 | 66 (57, 73) | 63 (56, 69) | <0.001 |
| **Region name** | 19,692 |  |  | <0.001 |
| East Midlands |  | 729 (9.5%) | 1,135 (9.4%) |  |
| East of England |  | 1,710 (22%) | 2,587 (21%) |  |
| London |  | 985 (13%) | 1,132 (9.4%) |  |
| North East |  | 308 (4.0%) | 660 (5.5%) |  |
| North West |  | 853 (11%) | 1,274 (11%) |  |
| South East |  | 1,472 (19%) | 2,421 (20%) |  |
| South West |  | 621 (8.1%) | 1,068 (8.9%) |  |
| Wales |  | 184 (2.4%) | 293 (2.4%) |  |
| West Midlands |  | 421 (5.5%) | 760 (6.3%) |  |
| Yorkshire and The Humber |  | 373 (4.9%) | 706 (5.9%) |  |
| **IMD Quintile** | 19,692 |  |  | 0.5 |
| (Most Deprived) 1 |  | 551 (7.2%) | 909 (7.6%) |  |
| 2 |  | 1,046 (14%) | 1,648 (14%) |  |
| 3 |  | 1,607 (21%) | 2,444 (20%) |  |
| 4 |  | 1,981 (26%) | 3,203 (27%) |  |
| (Least Deprived) 5 |  | 2,471 (32%) | 3,832 (32%) |  |
| **Sex** | 19,692 |  |  | 0.2 |
| Male |  | 3,297 (43%) | 5,307 (44%) |  |
| Female |  | 4,359 (57%) | 6,729 (56%) |  |
| **Ethnicity** | 19,692 |  |  |  |
| White British |  | 6,842 (89%) | 11,007 (91%) |  |
| White Irish |  | 128 (1.7%) | 170 (1.4%) |  |
| White Other |  | 341 (4.5%) | 457 (3.8%) |  |
| Mixed |  | 67 (0.9%) | 91 (0.8%) |  |
| South Asian |  | 164 (2.1%) | 171 (1.4%) |  |
| Other Asian |  | 54 (0.7%) | 61 (0.5%) |  |
| Black |  | 29 (0.4%) | 42 (0.3%) |  |
| Other Ethnicity |  | 31 (0.4%) | 37 (0.3%) |  |
| **Clinical Vulnerability** | 19,692 |  |  | <0.001 |
| Clinically extremely vulnerable |  | 1,095 (14%) | 1,413 (12%) |  |
| Clinically vulnerable |  | 2,424 (32%) | 3,450 (29%) |  |
| No sign of clinical vulnerability |  | 4,137 (54%) | 7,173 (60%) |  |
| **Followup Duration** | 19,692 | 83 (65, 100) | 67 (50, 83) | <0.001 |
| **SARs-CoV-2 infection** | 19,692 | 582 (7.6%) | 864 (7.2%) | 0.3 |
| <sup>1</sup> Median (IQR); n (%) |  |  |  |  |
| <sup>2</sup> Wilcoxon rank sum test; Pearson's Chi-squared test; Fisher's exact test |  |  |  |  |

Both groups were followed up to a maximum of 129 days (from September 16th until January 23^rd^, 2022), with ChAdOx1 individuals producing a median follow-up duration of 67 days (IQR: 50, 83), whilst BNT162b2 had a median follow-up duration of 83 days (IQR: 65, 100). At 129 days, ChAdOx1 participants experienced an incidence of 71.8 infections per 1,000 (95%CI: 67.2 to 76.5) vaccinated individuals whilst BNT162b2 participants experienced an incidence of 76 infections per 1,000 (95%CI: 70.2 to 82.2) vaccinated individuals.

### Crude Analysis

ChAdOx1 produces a pooled unadjusted hazard ratio of 0.95 [95% HR: 0.84-1.06] in the incidence of SARS-CoV-2 infection when compared to BNT162b2 at up to 129 days. For the same period, the pooled unadjusted hazard ratio for females (reference being male) was 0.95 [95%CI: HR: 0.85 - 1.05]. Per year of age, the hazard ratio was 0.97 [95%CI: HR: 0.96 - 0.97]. With the reference variable being “No sign of clinical vulnerability”, the pooled unadjusted hazard ratio for clinically vulnerable was 0.90 [95%: HR: 0.77 - 1.06] and 0.96 [95%: HR: 0.80 - 1.15] for “Clinically extremely vulnerable”. The least deprived groups had a protective effect against SARs-CoV-2 infections compared to the most deprived groups. See Figure 2 for unadjusted hazard ratio estimates.

**Figure 2:**
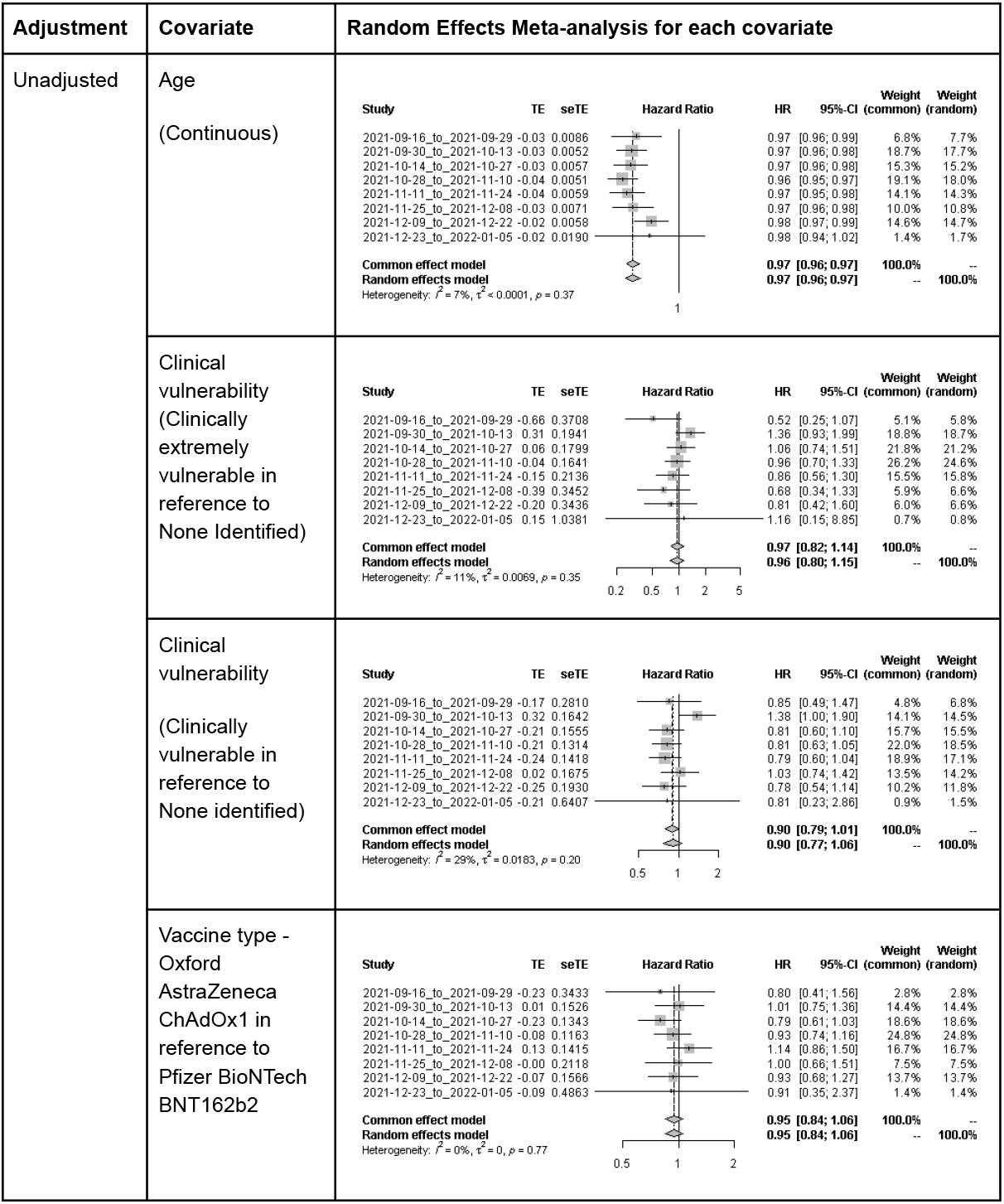
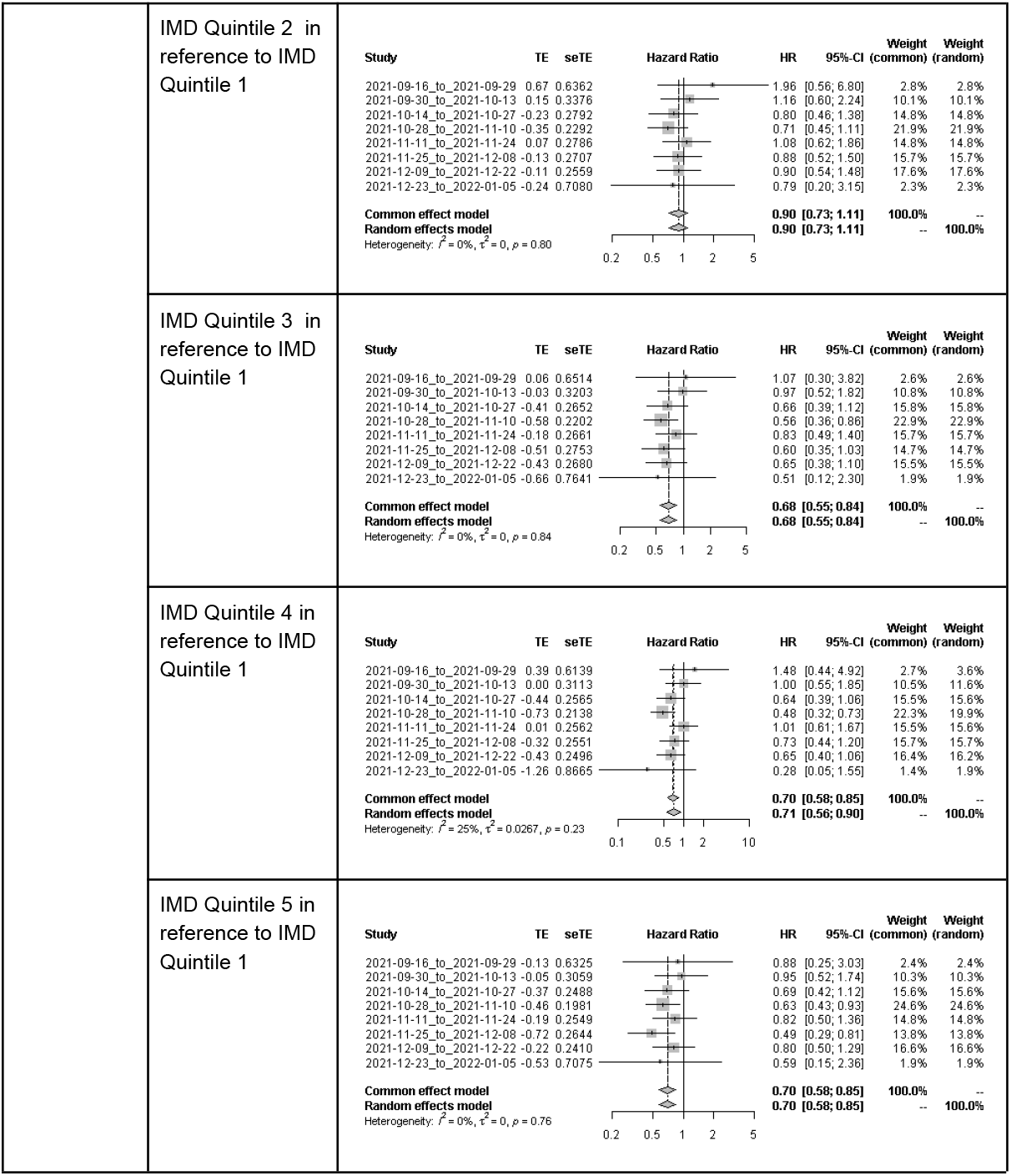
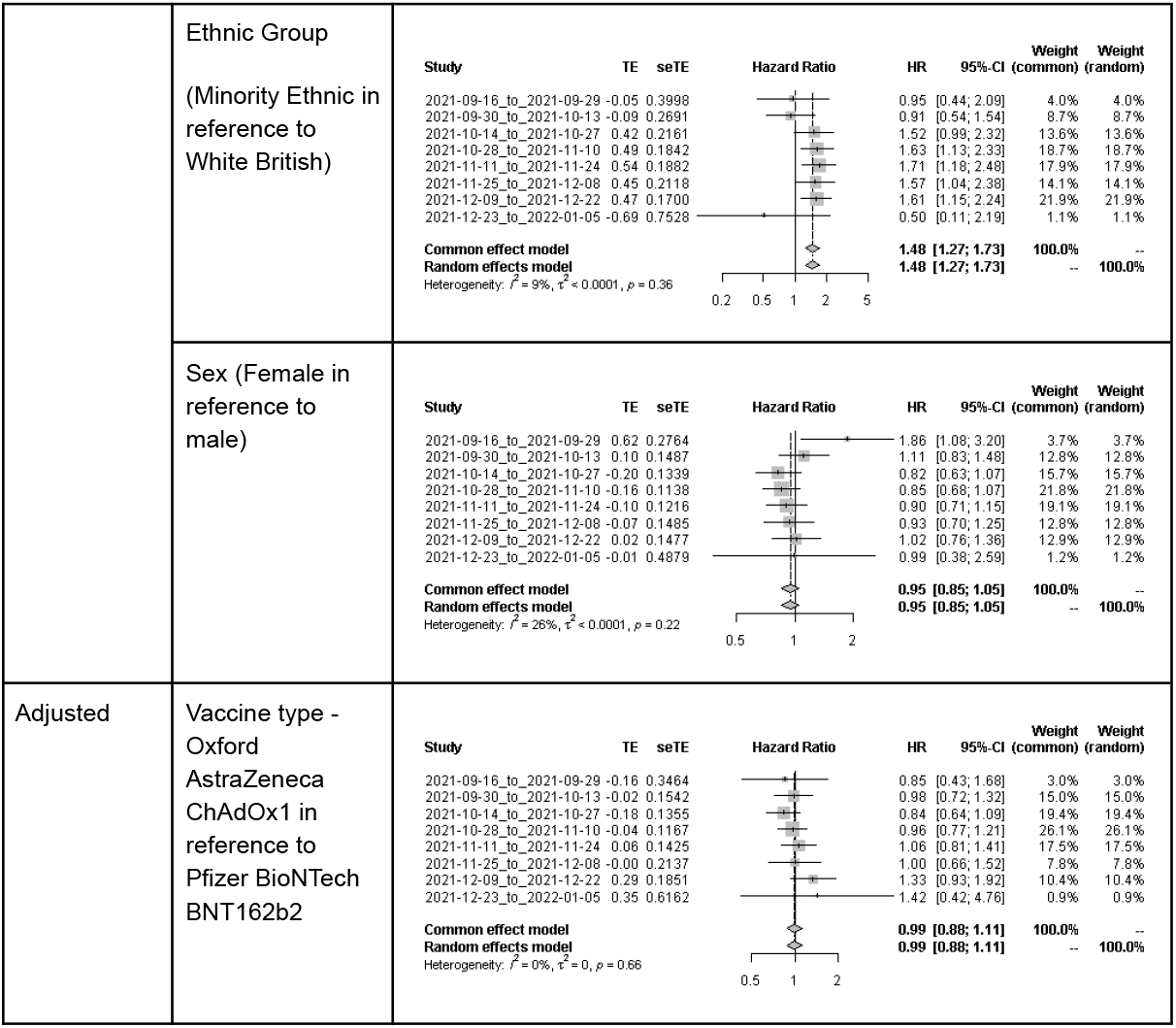
Adjusted and unadjusted random-effects meta-analysis for each covariate.

The change in rates of cumulative infections changed for all cohorts since the Omicron variant became the dominant strain of SARs-CoV-2 in the UK; those who were vaccinated earlier had a longer period of cumulative stability compared to those who were vaccinated closer or after the Omicron was declared the dominant SARs-CoV-2 strain in the UK. See Figure 3 for the cumulative incidence rate of SARs-COV-2 infections grouped by vaccination cohort.

**Figure 3:**
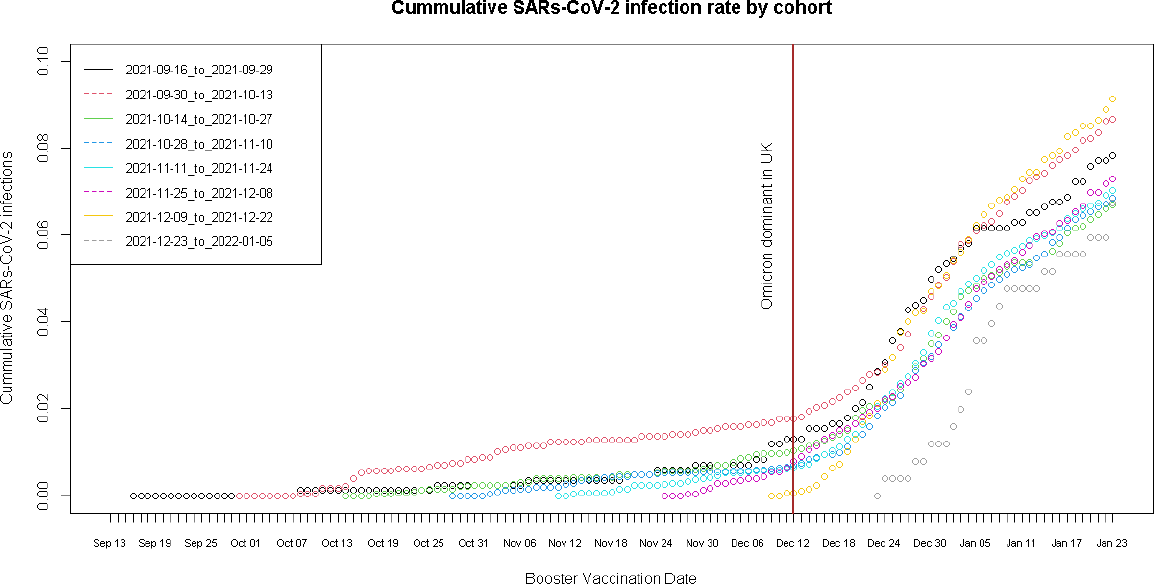
Cumulative SARs-CoV-2 incidence rate by vaccination date cohort. Between the date of booster vaccination and end of study, the rates of increase in cumulative proportions by cohort were: Cohort: 2021-09-16 to 2021-09-29: 0.055 [95%CI: 0.049 - 0.062] percent increase per day Cohort: 2021-09-30 to 2021-10-13: 0.066 [95%CI: 0.060 - 0.073] percent increase per day Cohort: 2021-10-14 to 2021-10-27: 0.064 [95%CI: 0.057 - 0.071] percent increase per day Cohort: 2021-10-28 to 2021-11-10: 0.078 [95%CI: 0.070 - 0.086] percent increase per day Cohort: 2021-11-11 to 2021-11-24: 0.107 [95%CI: 0.098 - 0.098] percent increase per day Cohort: 2021-11-25 to 2021-12-08: 0.136 [95%CI: 0.129 - 0.143] percent increase per day Cohort: 2021-12-09 to 2021-12-22: 0.231 [95%CI: 0.220 - 0.241] percent increase per day Cohort: 2021-12-13 to 2022-01-05: 0.227 [95%CI: 0.207 - 0.246] percent increase per day

### Adjusted Analysis

After adjusting for age at vaccination, clinical vulnerability, IMD quintile, minority ethnic status and sex, the pooled adjusted hazard ratio for ChAdOx1 was 0.99 [95%CI: aHR: 0.88 - 1.11] suggesting no significant difference in vaccination effectiveness when compared to BNT162b2 after receiving an mRNA based booster vaccine (see Figure 2 – adjusted section)

### Sensitivity Analysis

Whilst the primary analysis only analysed vaccine induced protection, such results are less likely to be pragmatic due to the increased proportions of those being infected by SARs-CoV-2. Therefore, we conducted a sensitivity analysis that included those with a prior SARs-CoV-2 infection to their booster dose but we adjusted for this in our modelling to account for the impact of natural infection on the protection provided by the nucleocapsid antibody. This increased our cohort from 19,692 participants to 22,062 individuals with 8,472 receiving BNT162b2 and 13,590 receiving ChAdOx1 as their primary vaccine courses. Socio-demographics characteristics remained similar to the primary analysis (See supplementary material 2). Both the univariable and multivariable models remained similar to the primary analysis; having a prior SARs-CoV-2 infection prior to the booster vaccine demonstrated a protective effect in both univariable (HR: 0.72 [95%CI: 0.60 - 0.87]) and multivariable model (HR: 0.65 [95CI: 0.55 - 0.77]) (See supplementary material 3).

## Discussion

Our analysis was conducted in a community cohort of 19,692 people across England and Wales who received their booster vaccination between September 16^th^ 2021 and January 5^th^ 2022. We followed people up for risk of SARS-CoV-2 infection between September 16^th^ 2021 and January 23^rd^ 2022 and found that people who received ChAdOx1 vaccinations as their primary course had no difference in the incidence of SARS-CoV-2 infection compared to BNT162b2 during follow-up after we accounted for differences in demographic and clinical characteristics between our comparison groups as well as the time of vaccination.

Our analysis used a community sample design from across England and Wales, in cohort with diversity in terms of age, sex, and geographical location. We estimated effectiveness in a cohort with a median follow up of two months after a booster vaccination, and the majority of infections occurred during a period when Omicron became the dominant variant in the UK. A particular strength of our analysis was our ability to estimate vaccine effectiveness in a cohort that included large numbers of people who were either clinical vulnerable or clinically extremely vulnerable – a group that was prioritised for booster doses based upon need. Using this sample, we applied a trial emulation framework to mitigate against confounding by indication. As a result of this study design, our results are more likely to reflect a randomised controlled trial evaluating the same question.

Our staggered cohort approach has additional strengths. First, it enables us to account for the demographic and clinical risk factors of vaccines and make comparisons between similar demographically and clinically similar groups; this was demonstrated in the changing median age and declining clinical vulnerable status of our cohorts. Second, our approach helps control for changes in SARS-CoV-2 transmission rates driven by changes in public health policy such as the vaccination efforts (e.g., prioritised distribution), mask usage, limitations on movement as well the emergence of new SARS-CoV-2 variants and their transition to becoming the dominant SARs-CoV-2 strain into England and Wales which we graphically demonstrated in Figure 3. Therefore our approach controls for measured time-varying confounders and to some extent, it goes some way to mitigating against the impact of unmeasured time-varying confounders (i.e. SARs-CoV-2 strain).

Due to the reliance on self-reported observational studies, there is a risk of inconsistent and inaccurate data recording; however, this was mitigated through linkage to external data sources such as SGSS to complement missing incidence SARS-CoV-2 infections and NIMs to complement missing vaccination data. We measured the risk of SARS-CoV-2 infection as our primary outcome, and whilst this precedes hospitalisation or death, we were not able to look at these more severe outcomes, which is a limitation of our study. Our use of observational data may mean that there is residual and uncontrolled confounding. Unlike test-negative designs our approach does not implicitly control for differences in testing behaviour between groups, but since we are comparing vaccine regimes rather than vaccinated and unvaccinated individuals we do not expect confounding by differential testing behaviour. Using multiple staggered cohorts reduces each cohort size, and as a result, we had difficulties with analysing certain covariates such as geographical region and ethnicity, which we had to combine into an aggregated category. We did not include occupation or geographical risk in our analyses, and these may result in imbalances in the comparison arms as both risks of exposure to SARS-CoV-2 infection and access to BNT162b2 varied geographically (due to its cold storage requirements) and by occupation (e.g., health and social care workers).

We found evidence of the same effectiveness of BNT162b2 compared to ChAdOx1 vaccines against SARS-CoV-2 infection after receiving a booster vaccination in England and Wales, a finding that contrasts previous analysis showing that prior to such boosters those who had ChAdOx1 as their primary course were at higher risk of a SARs-CoV-2 infection. In other analyses we have demonstrated that antibody levels are substantially higher following a primary course of BNT162b2 than following a primary course of BNT162b2, that antibodies wane following a log-linear pattern and that risk of infection is increased in those with lower antibody levels. Thus we hypothesise that differential effectiveness of BNT162b2 and ChadOx1 primary courses against infection are related to different antibody levels. We have also shown that following an mRNA booster dose antibody levels are similar regardless of the primary course and hypothesise that this accounts for the similar effectiveness of mRNA boosters regardless of primary regime. Our findings demonstrate the importance of mRNA booster doses in maintaining protection, particularly for those with a primary course of ChadOx1.

## Data Availability

We aim to share aggregate data from this project on our website and via a “Findings so far” section on our website - https://ucl-virus-watch.net/. We will also be sharing individual record level data on a research data sharing service such as the Office of National Statistics Secure Research Service. In sharing the data we will work within the principles set out in the UKRI Guidance on best practice in the management of research data. Access to use of the data whilst research is being conducted will be managed by the Chief Investigators (ACH and RWA) in accordance with the principles set out in the UKRI guidance on best practices in the management of research data. We will put analysis code on publicly available repositories to enable their reuse.

## Funding

The Virus Watch study is supported by the MRC Grant Ref: MC_PC 19070 awarded to UCL on 30 March 2020 and MRC Grant Ref: MR/V028375/1 awarded on 17 August 2020. The study also received $15,000 of Facebook advertising credit to support a pilot social media recruitment campaign on 18th August 2020. This study was also supported by the Wellcome Trust through a Wellcome Clinical Research Career Development Fellowship to RA [206602]. SB and TB are supported by an MRC doctoral studentship (MR/N013867/1). The funders had no role in study design, data collection, analysis and interpretation, in the writing of this report, or in the decision to submit the paper for publication.

## Data Availability

We aim to share aggregate data from this project on our website and via a “Findings so far” section on our website - https://ucl-virus-watch.net/. We will also be sharing individual record level data on a research data sharing service such as the Office of National Statistics Secure Research Service. In sharing the data we will work within the principles set out in the UKRI Guidance on best practice in the management of research data. Access to use of the data whilst research is being conducted will be managed by the Chief Investigators (ACH and RWA) in accordance with the principles set out in the UKRI guidance on best practice in the management of research data. We will put analysis code on publicly available repositories to enable their reuse.

## Contributors

Study Conceptualisation: VGN, AY, RWA

Formal analysis: VGN, AY, RWA

Project administration: JK, VGN, SB, TB, AMDN, MS, SW.

Data curation: VGN, AMDN, CY, WLEF, MS.

Writing (original draft preparation): VGN, AY, RWA, ACH.

Writing (review and editing): All authors.

Data Access: All authors had full access to the data used in the study.

## Supplementary Material

**Supplementary Figure 1:**
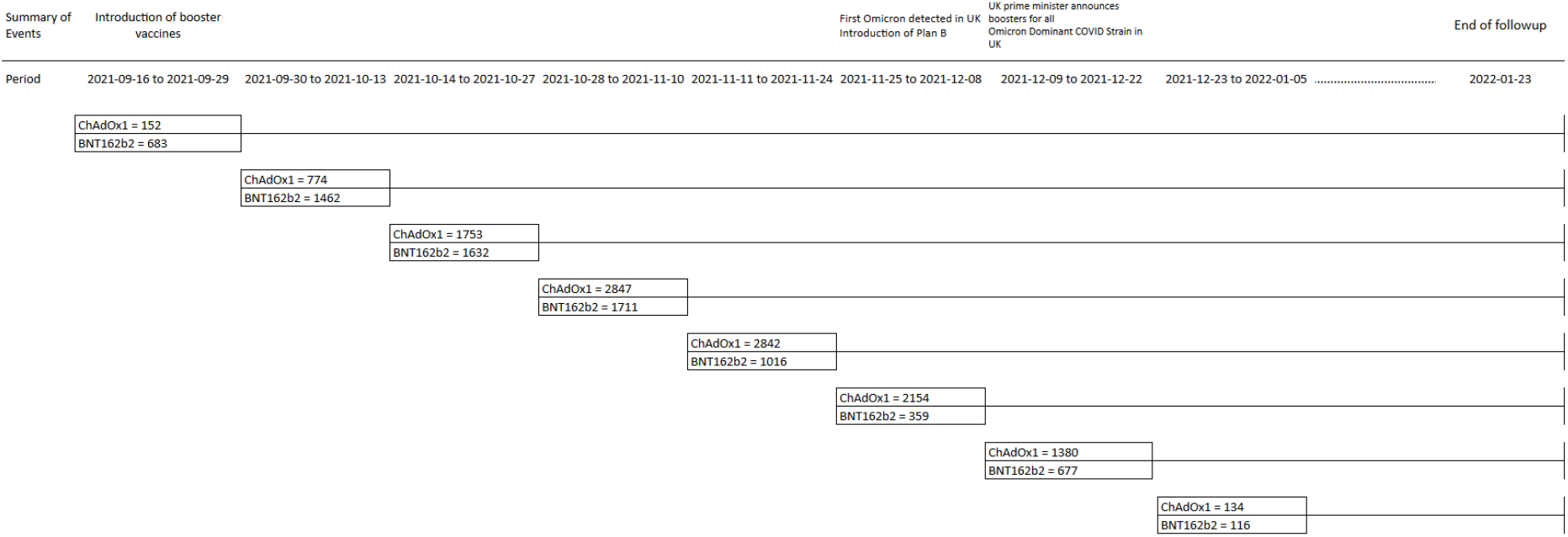
Cohort recruitment diagram through time, including a description of time varying events

**Supplementary Material 2:**
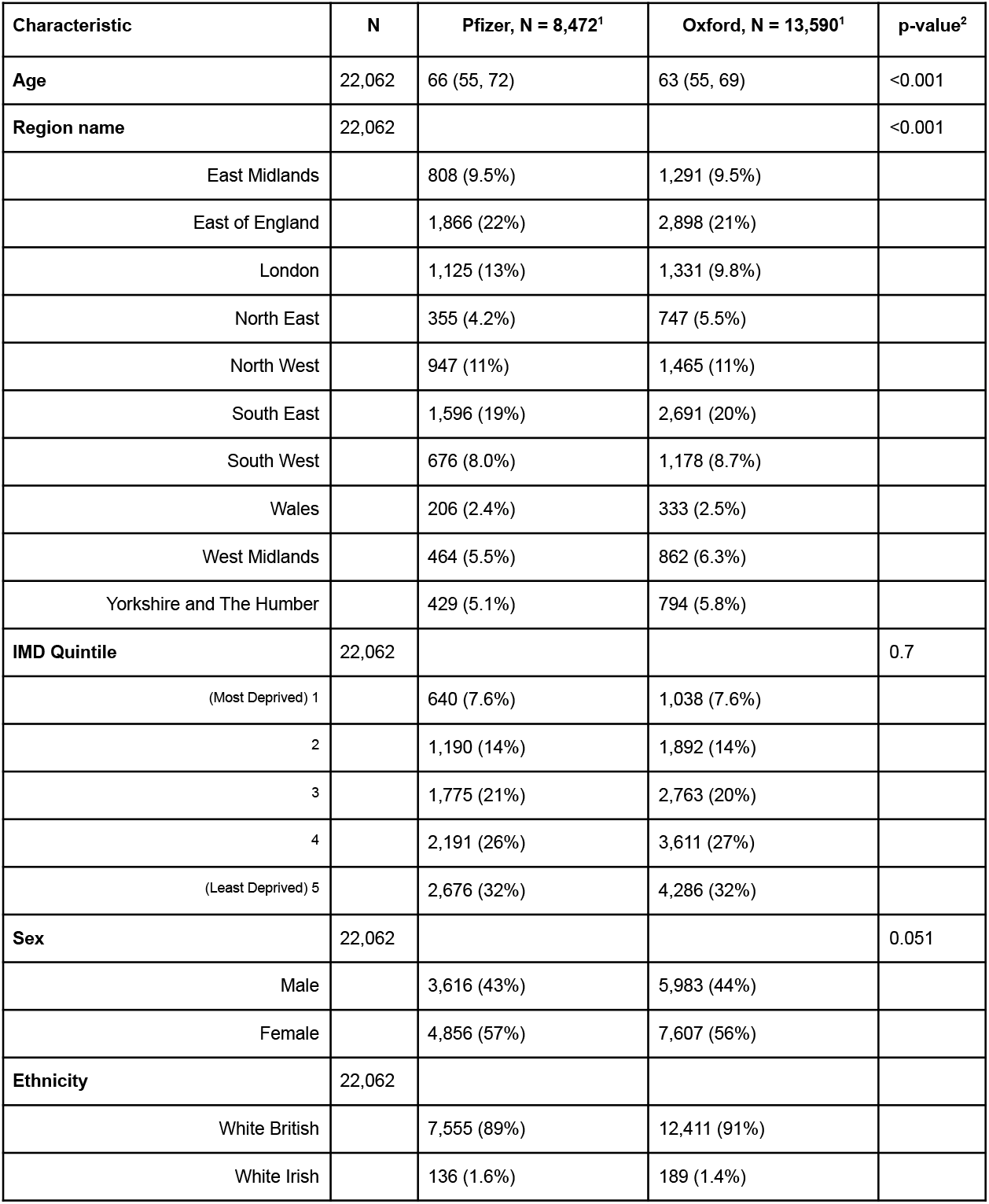
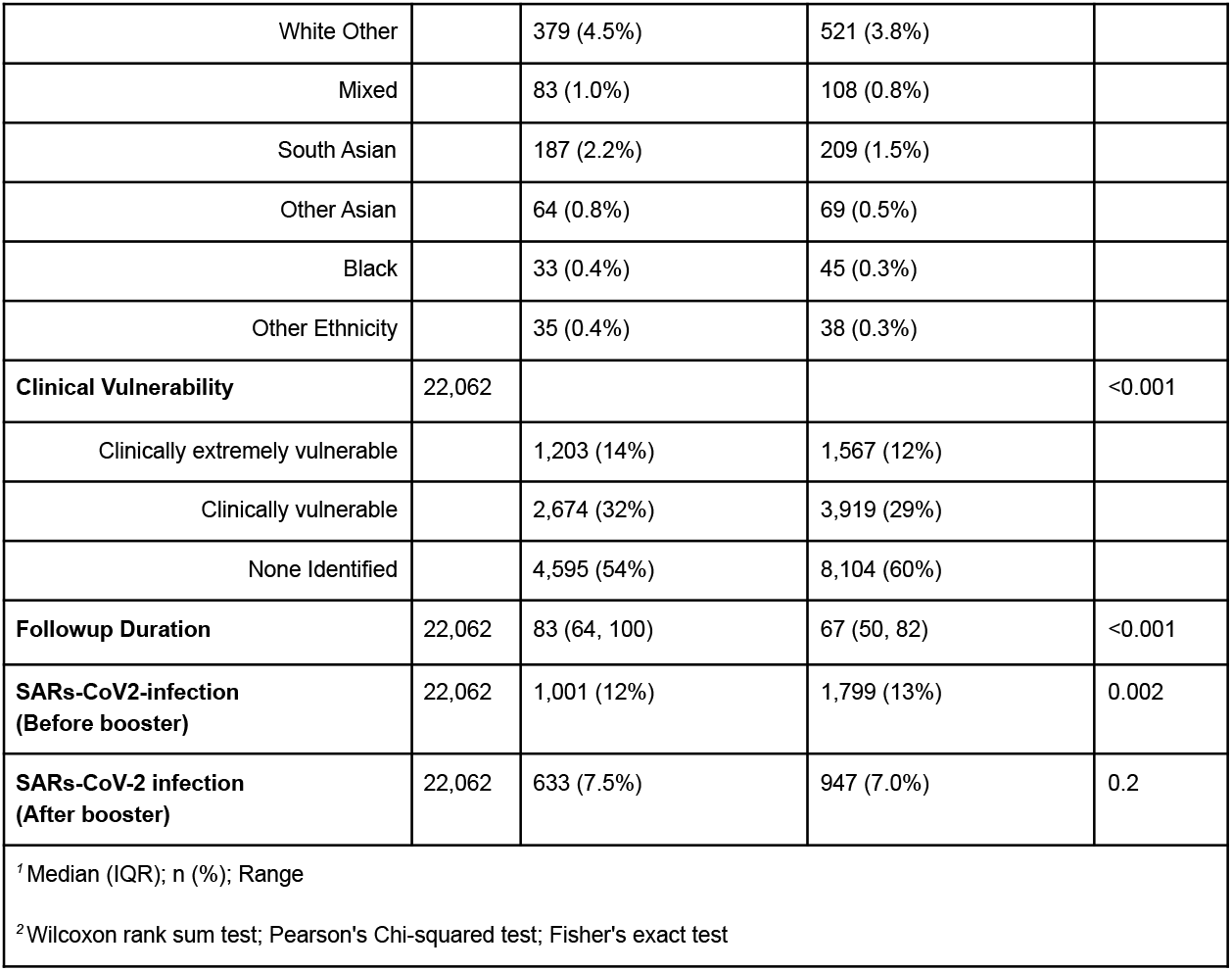
Demographics breakdown of the sensitivity analysis

**Supplementary Material 3.**
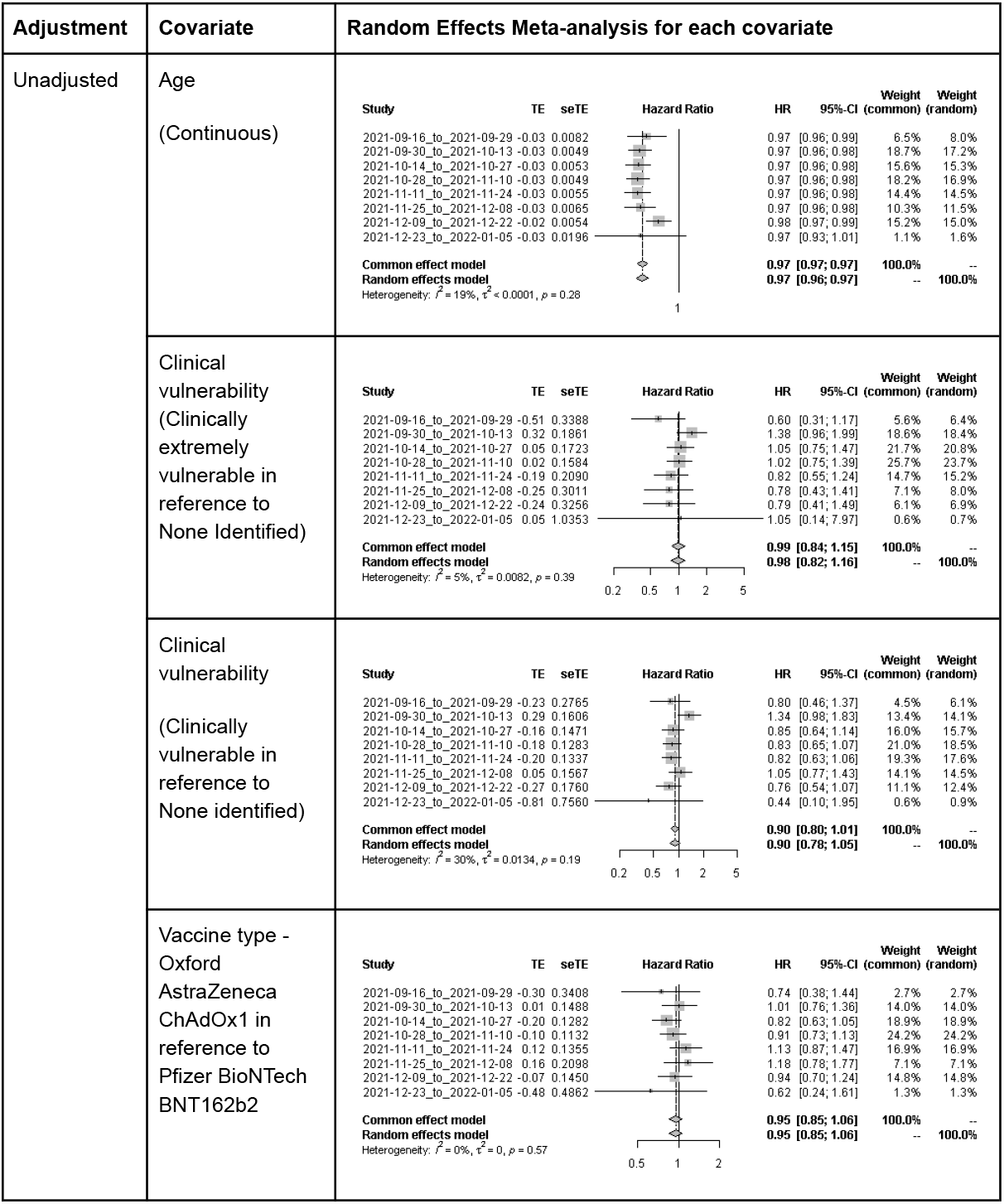
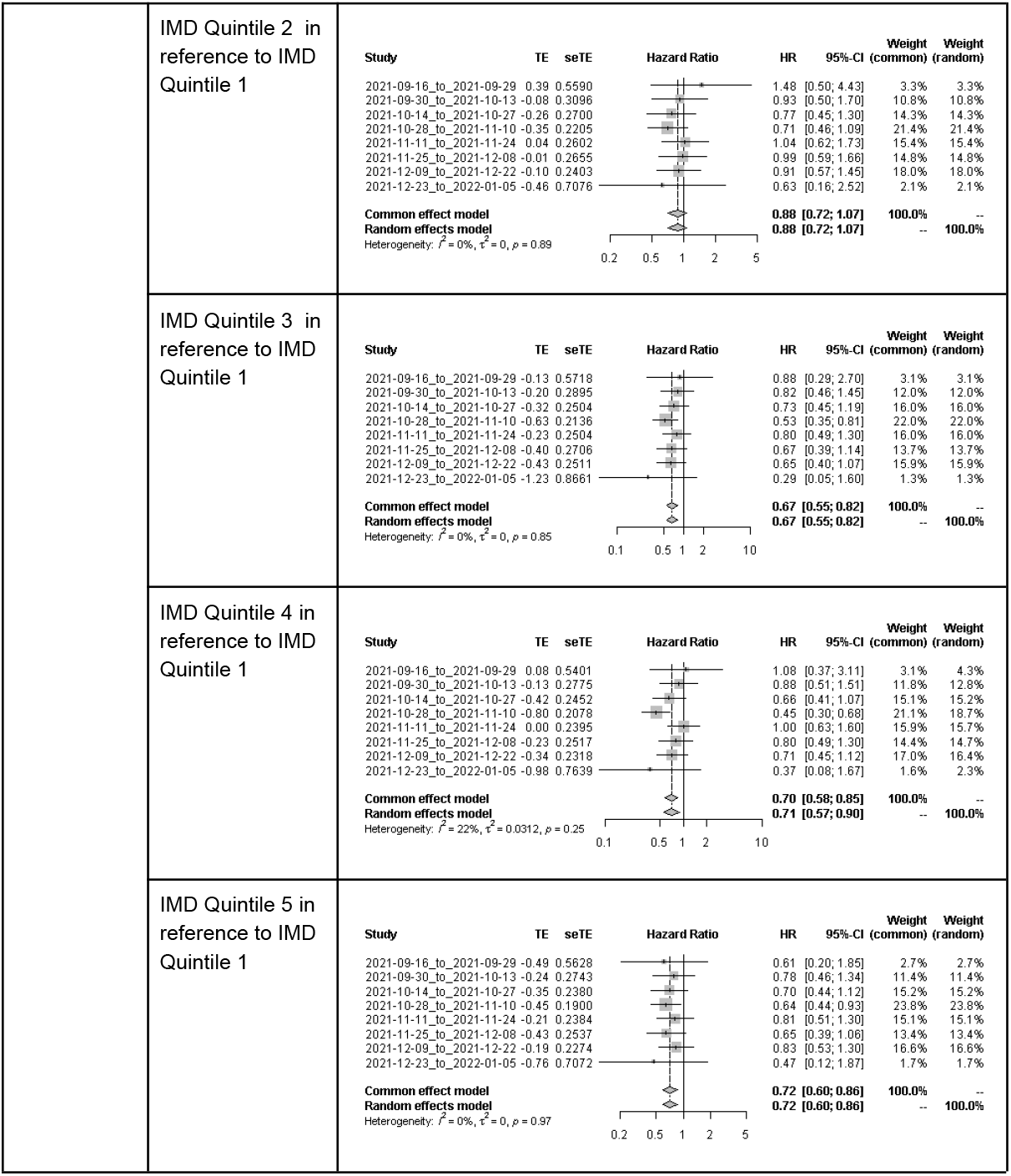
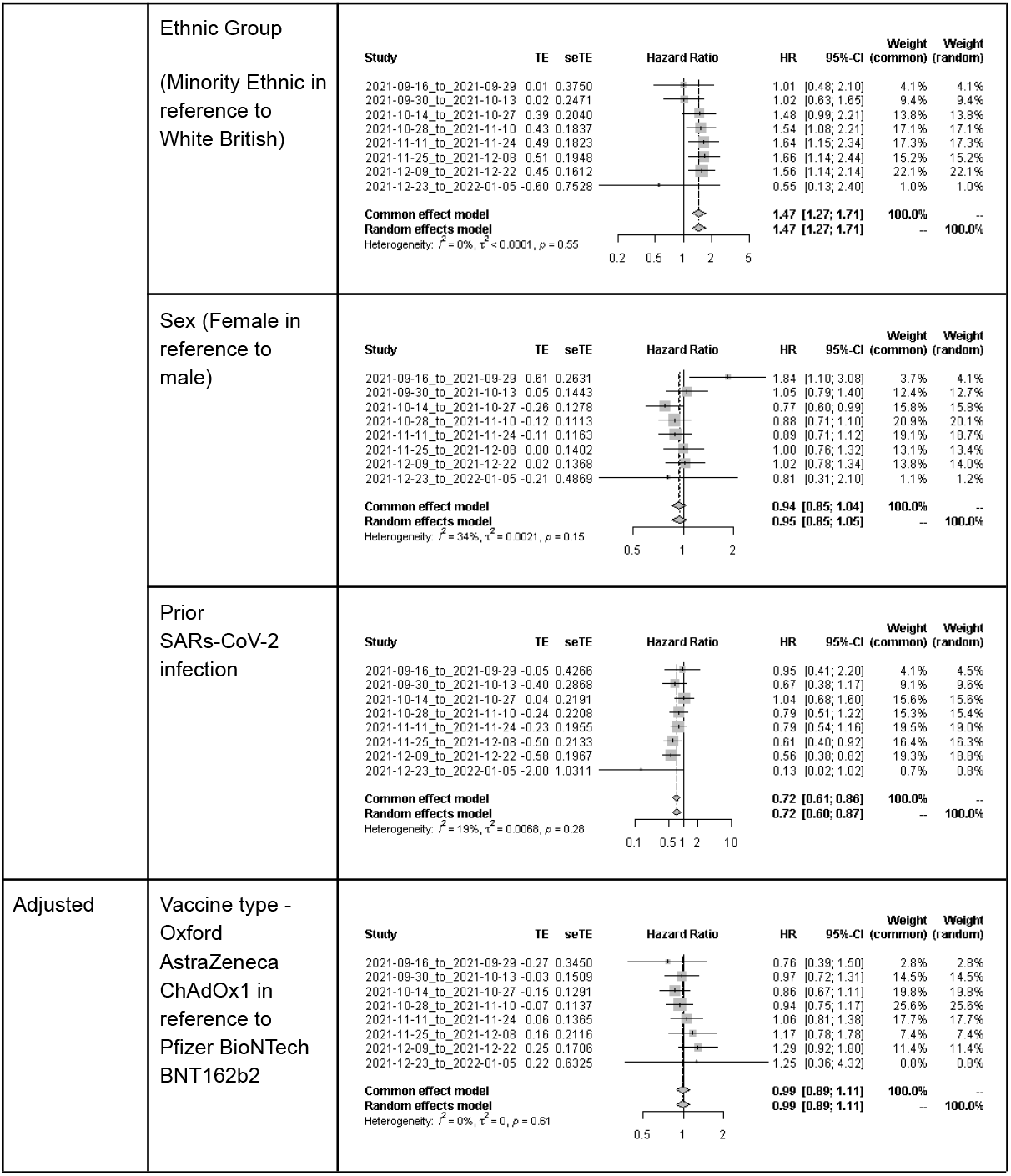
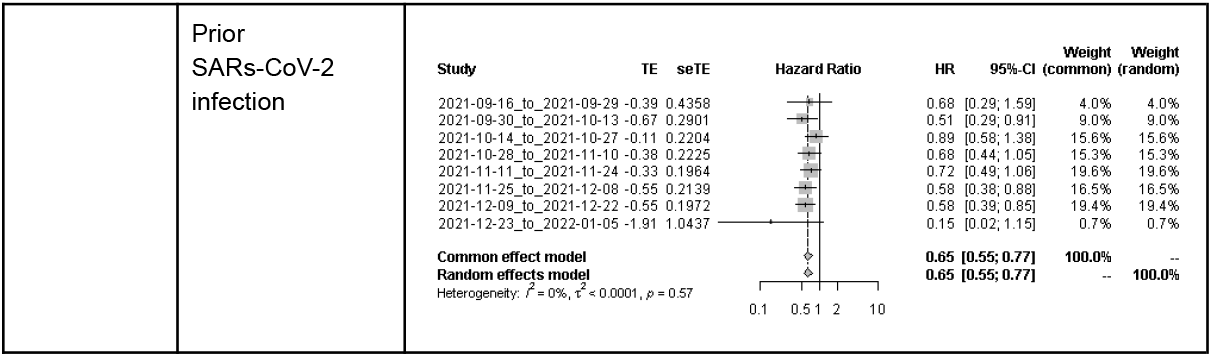
Adjusted and unadjusted random-effects meta-analysis for each covariate for the sensitivity analysis

## Notes

### Competing Interest Statement

ACH serves on the UK New and Emerging Respiratory Virus Threats Advisory Group.

## References

1. Nguyen VG, Yavlinsky A, Beale S, et al. Comparative Effectiveness of ChAdOx1 versus BNT162b2 Vaccines against SARS-CoV-2 Infections in England and Wales: A Cohort Analysis Using Trial Emulation in the Virus Watch Community Data. Epidemiology; 2021. doi:10.1101/2021.12.21.21268214

2. Shrotri M, Navaratnam AMD, Nguyen V, et al. Spike-antibody waning after second dose of BNT162b2 or ChAdOx1. The Lancet. 2021;398(10298):385–387. doi:10.1016/S0140-6736(21)01642-1

3. Munro APS, Janani L, Cornelius V, et al. Safety and immunogenicity of seven COVID-19 vaccines as a third dose (booster) following two doses of ChAdOx1 nCov-19 or BNT162b2 in the UK (COV-BOOST): a blinded, multicentre, randomised, controlled, phase 2 trial. The Lancet. 2021;398(10318):2258–2276. doi:10.1016/S0140-6736(21)02717-3

4. Andrews N, Stowe J, Kirsebom F, et al. Effectiveness of COVID-19 Vaccines against the Omicron (B.1.1.529) Variant of Concern. Epidemiology; 2021. doi:10.1101/2021.12.14.21267615

5. Ozasa K, Fukushima W. Commentary: Test-Negative Design Reduces Confounding by Healthcare-Seeking Attitude in Case-Control Studies. J Epidemiol. 2019;29(8):279–281. doi:10.2188/jea.JE20180177

6. Dean NE, Halloran ME, Longini, Jr IM. Temporal Confounding in the Test-Negative Design. Am J Epidemiol. 2020;189(11):1402–1407. doi:10.1093/aje/kwaa084

7. Hernán MA, Alonso A, Logan R, et al. Observational Studies Analyzed Like Randomized Experiments: An Application to Postmenopausal Hormone Therapy and Coronary Heart Disease. Epidemiology. 2008;19(6):766–779. doi:10.1097/EDE.0b013e3181875e61

8. Hayward A, Fragaszy E, Kovar J, et al. Risk Factors, Symptom Reporting, Healthcare-Seeking Behaviour and Adherence to Public Health Guidance: Protocol for Virus Watch, a Prospective Community Cohort Study. Infectious Diseases (except HIV/AIDS); 2020. doi:10.1101/2020.12.15.20248254

9. Hernán MA, Robins JM. Using Big Data to Emulate a Target Trial When a Randomized Trial Is Not Available: Table 1. Am J Epidemiol. 2016;183(8):758–764. doi:10.1093/aje/kwv254

